# *In Vivo* Mitochondrial ATP Production Is Improved in Older Adult Skeletal Muscle After a Single Dose of Elamipretide in a Randomized Trial

**DOI:** 10.1101/2020.09.30.20200493

**Authors:** Baback Roshanravan, Sophia Z. Liu, Eric G. Shankland, John K. Amory, H. Thomas Robertson, David J. Marcinek, Kevin E. Conley

## Abstract

**Background:** Loss of mitochondrial function contributes to fatigue, exercise intolerance and muscle weakness, and is a key factor in the disability that develops with age and a wide variety of chronic disorders. Here, we describe the impact of a first-in-class cardiolipin-binding compound that is targeted to mitochondria and improves oxidative phosphorylation capacity (Elamipretide, ELAM) in a randomized, double-blind, placebo-controlled clinical trial.

**Methods:** Non-invasive magnetic resonance and optical spectroscopy provided measures of mitochondrial capacity (ATP_max_) with exercise and mitochondrial coupling (ATP supply per O_2_ uptake; P/O) at rest. The first dorsal interosseous (FDI) muscle was studied in 39 healthy older adult subjects (60 to 85 yrs of age; 46% female) who were enrolled based on the presence of poorly functioning mitochondria. We measured volitional fatigue resistance by force-time integral over repetitive muscle contractions.

**Results:** A single ELAM dose elevated mitochondrial energetic capacity *in vivo* relative to placebo (ΔATP_max_; *P*=0.055, %ΔATP_max_; *P*=0.045) immediately after a 2-hour infusion. No difference was found on day 7 after treatment, which is consistent with the half-life of ELAM in human blood. No significant changes were found in resting muscle mitochondrial coupling. Despite the increase in ATP_max_ there was no significant effect of treatment on fatigue resistance in the FDI.

**Conclusions:** These results highlight that ELAM rapidly and reversibly elevates mitochondrial capacity after a single dose. This response represents the first demonstration of a pharmacological intervention that can reverse mitochondrial dysfunction *in vivo* immediately after treatment in aging human muscle.

---

Age and disease are associated with a debilitating loss of function that leads to disability [1, 2], underlies morbidity [3] and has a substantial economic impact on society, including the healthcare system [4]. Mitochondrial dysfunction is a key part of the pathophysiology and reduced resilience associated with aging [5] and chronic disease [6] in multiple organ systems. Mitochondrial dysfunction is also associated sarcopenia and impaired physical endurance across multiple populations[7, 8]. This loss of muscle function is an important contributor to reduced quality of life and increased morbidity with age [9-11]. Despite observational data indicating the important role of mitochondria in aging and disease there are no approved treatments that directly target dysfunctional mitochondria in skeletal muscle. A new approach has emerged that targets dysfunctional lipids, involves repair rather than replacement of deteriorated components [12, 13], and acts rapidly after a single treatment to reverse dysfunction [14].

Elamipretide (ELAM) is a first-in-class compound that binds reversibly to the phospholipid cardiolipin (CL) located within the inner mitochondrial membrane to stabilizes its structure, promote cristae formation and curvature, and positively alters the electrostatic environment [15, 16] [17, 18]. Early studies inaccurately described ELAM as having antioxidant properties, [19-21] although more recent work has demonstrated that ELAM actually reduces production of superoxide and H_2_O_2_ particularly where overproduction resulted from dysfunctional mitochondria [22, 23]. Studies in isolated mitochondria, cells and intact organs reveal improved ETC flux, reduced mitochondrial generation of reactive oxygen species (ROS; H_2_O_2_), and elevated ATP generation with ELAM treatment after ischemic insult and heart failure [16, 24, 25]. In skeletal muscle, treatment with ELAM prevented the increase in mitochondrial oxidative stress and muscle wasting associated with disuse atrophy in mouse hindlimb and diaphragm. In aging mice a single IP injection of ELAM elevated the capacity to generate ATP, improved the coupling of oxidative phosphorylation (P/O) in vivo, and increased fatigue resistance in the tibialis anterior in old hindlimb muscle without affecting mitochondrial energetics in young muscle [14]. Running endurance was also elevated in the old mice after 8 days of daily injections of ELAM. More recently, an 8-week treatment with ELAM demonstrated similar increases in *in vivo* mitochondrial energetics and fatigue resistance in old mice, as well as a reversal of age-related redox stress[26]. It is noteworthy that 4 month treatment of aged mice with ELAM reduced mtROS emission and improved mitochondrial integrity in muscles from similar to the 8 week treatment, but did not demonstrate a reduction in fiber atrophy nor improvements in force and fatigue resistance in isolated muscles[26, 27], suggesting that improvements in performance with ELAM treatment may be potentiated by systemic effects beyond the myofiber fiber level. In a Phase 2 study 5 daily doses of ELAM increased the walking distance over 6 minutes (Six-Minute Walk Test [6MWT]) ELAM in individuals with a genetic mitochondrial myopathy [28], although the subsequent expanded Phase 3 trial failed to detect a change in the 6 MWT. Thus, studies from both mouse models and human disease indicate that both a better understanding of the effect of ELAM on in vivo mitochondrial energetics and the link between mitochondrial energetics and skeletal muscle function is necessary. Despite the trials focused on mitochondrial myopathy and other human trials in heart disease, ischemia reperfusion, and ongoing trials in Barth and mitochondrial associated vision disorders, there has been no direct measurement of mitochondrial function in a human clinical trial to determine if the mechanism of action of ELAM is mitochondrial in origin.

Here, we report the results from a randomized, double-blind, placebo controlled clinical trial that evaluated the safety and efficacy of a single dose of ELAM on surrogate endpoints of mitochondrial and muscle function in healthy older adults. Participants were selected on the basis of having evidence of impaired in vivo muscle mitochondrial energetics determined by dynamic, in vivo ^31^Phosphorus Magnetic Resonance Spectroscopy using a using a standardized protocol [29, 30]. The effects on the first dorsal interosseous (FDI) muscle of the hand was investigated for two reasons. First, handgrip strength is an important clinical measure of frailty and disease burden strongly associated with adverse health related outcomes across populations[31-34]. Second, this particular muscle shows clear changes in mitochondrial function with age [29] providing direct measurement of both mitochondrial energetics and muscle function (Supplementary Figure S3). We hypothesized that ELAM infusion will lead to immediate improvements in our primary surrogate endpoint, the maximal rate of ATP production (ATP_max_)immediately after a 2-hour infusion of ELAM.. Two secondary outcomes were evaluated: 1) the resting state coupling of oxidative phosphorylation (ATP/O_2_) to evaluate mechanisms of improvement and 2) exercise tolerance as evaluated by muscle performance after ELAM treatment. Evidence of efficacy of mitochondrial targeted therapeutics to improve muscle metabolism and function may guide future therapeutic development and inform pharmacologic or lifestyle interventions targeting improvements in quality of life in the older adults.

## METHODS

### Study Design and Participants

This randomized, double-blind, placebo-controlled trial was conducted at the University of Washington Medical Center. Adults 60 to 85 years of age were recruited through public lectures, mailers, and posted advertisements. Eligible participants were selected based on two criteria found to be linked to low muscle function: 1) muscle mitochondrial ATP synthesis capacity (ATP_max_<0.7 mM/sec) and 2) the coupling of oxidative phosphorylation (ATP synthesized per O_2_ uptake ÷ 2 < 1.9, [P/O]) [29, 30]. These fluxes were assessed by *in vivo* magnetic resonance and optical spectroscopy (**Supplementary Figures S1 and S2**) [29, 30]. Inclusion and exclusion criteria are listed in **Supplementary Tables S1 and S2**. All procedures and protocols were in accordance with the tenets of the declaration of Helsinki. The study protocol was approved by the University of Washington Human Subjects Division and Western Institutional Review Board (IRB00000533). All subjects provided written informed consent before entering the trial. The study was registered with ClinicalTrials.gov (identifier: NCT02245620).

### Power Calculations, Randomization, and Masking

The group size of 20 subjects each for treatment and placebo was based on a mean change in P/O of 0.4 and standard deviation of ± 0.38 to provide a power of 0.9 at the 0.05 level of significance in healthy subjects 65 to 80 years of age[35]. An amended protocol placed ATP_max_ as the primary outcome and P/O as the secondary outcome. Randomization was performed at a 1:1 ratio by a computer-generated random sequence using an Interactive Web-Response System (IWRS). Both participants and study personnel were blinded to treatment assignment during the duration of the study. ELAM was administered intravenously for 2 hours at 0.25 mg**/**kg body weight per hour, which is the level found to be both safe and effective in two human dose-escalation studies [28, 36]. Placebo consisted of the vehicle used for the active drug, which contained preservative level concentrations of trehalose dihydrate, glutathione, and sodium hydroxide/acetic acid in physiological saline.

### Outcomes

The primary endpoint was the change in *in vivo* skeletal muscle mitochondrial capacity (ATP_max_) with exercise after 2 hours of infusion compared to baseline. The secondary endpoints were resting mitochondrial coupling (P/O) and exercise tolerance (force-time integral). The coupling of phosphorylation (ATP supply) to oxidation (O_2_ uptake) by the mitochondria was expressed as P/O (ATP/O_2_ ÷ 2). The change in muscle performance was measured as the sum of the muscle force (force-time integral, FTI) normalized to the maximum voluntary contraction (MVC) generated during voluntary contractions. This protocol involved step increases of contraction frequency starting at 60 contractions per minute [cpm] and increasing frequency 10 cpm each minute until exhaustion. Primary and secondary endpoints were reassessed 7 days post-infusion.

### Study Procedures

The study consisted of 4 visits. A screening visit (Visit 1 or V1) determined eligibility and included a general health exam including ECG, blood tests, and non-invasive measures of muscle mitochondrial energetics and exercise tolerance of the hand muscle (first dorsal interosseous [FDI], **Supplementary Figure S3**). Those found to qualify for the study returned within 28 days of V1, were infused with either placebo or drug, and all procedures were repeated immediately following infusion (Visit 2 or V2). The exercise tolerance test was repeated a day after the infusion (Visit 3 or V3). All procedures were repeated one week after infusion (Visit 4 or V4).

### MRS/OS Protocol

The protocol for the phosphorus magnetic resonance spectroscopy (^31^P MRS) and optical spectroscopy (OS) has been previously published [29, 30]. These two spectroscopic measurements were made on the muscle during a period of ischemia. Probes for each measurement were placed at the same location on the skin surface of the FDI (**Supplementary Figure S3**). This allowed us to separate O_2_ uptake from ATP flux in resting muscle *in vivo* (**Supplementary Figure S2**). P/O is a measure of the efficiency of ATP generation in resting muscle (ATP/O_2_ ÷ 2). The mitochondrial oxidative phosphorylation capacity (ATP_max_) was determined as described [30, 37]. Briefly, a short exercise bout involving the index finger was performed for a period of 20 to 30 seconds so that PCr was reduced by ∼50%, while maintaining muscle pH>6.8. The PCr recovery was measured over 6 min to determine a time constant of recovery (t_PCr_) to yield ATP_max_ (= PCr_rest_/ t_PCr_]) where PCR_rest_ = 25.5 mM. The ATP_max_ is the standard for characterizing mitochondrial ATP capacity *in vivo* and is directly related both to mitochondrial markers of oxidative phosphorylation and mitochondrial oxidation *in vivo*.

### Muscle Performance Testing

Exercise tolerance was tested by measuring the sum of force generated by the FDI muscle during repeated isometric contractions until exhaustion (**Supplementary Figure S3**). The FDI is a model system for both energetic and mechanics measures [38] as well as mitochondrial aging [29]. The maximum voluntary contraction (MVC) was measured as the average of 3 maximum contractions separated by 5 sec and sustained for 3 sec each. The exercise level was set at 70% of the MVC and the exercise began at a frequency of 60 contractions per minute (cpm) for the first minute. This frequency was increased at a rate of 10 cpm with each minute until exhaustion. The FTI was measured as the sum of the force generated by these contractions during the test and normalized to the subject’s MVC.

### Statistical Analysis

The primary analysis set was the intent to treat study population. The primary endpoint test was the difference in the mean change from baseline to immediately after infusion between ELAM and the placebo group in a Student’s t-test in an analysis of covariance (ANCOVA) framework, with baseline as a covariate. The secondary endpoints (P/O and FTI) were tested as a mean change from baseline between treatment groups immediately after and 7 days post-infusion. Statistical tests were 2-sided and evaluated at the 0.05 level of significance. No adjustment for multiple comparisons was made per the pre-specified statistical plan.

## RESULTS

### Participant Characteristics

Thirty-nine subjects, from 120 screened potential participants, were randomized to participate in the study based on overall good general health but presenting with low mitochondrial function (ATP_max_ <0.7 mM sec^-1^ and P/O <1.9) (**Figure 1**). Forty-seven subjects [39%] were excluded with high mitochondrial function, while 34 subjects [28%] were excluded due to clinical criteria. ATPmax data was not available for V2 for 1 subject in both the PL and ELAM groups due to problems with data collection. An additional participant in the placebo group had an MRS result that did not meet our quality control criteria (motion artifact) after inclusion in the database; that scan was excluded from the analysis of the primary endpoint. A summary of the demographic and clinical assessments appears in **Table 1**.

**Table 1.** Baseline Characteristics of Study Population.

| **Table 1. Baseline Characteristics of Study Population** |  |  |
| --- | --- | --- |
|  | **Placebo**<br>**(n=20)** | **Elamipretide**<br>**(n=19)** |
| **Demographic** |  |  |
| Age (years, mean [±SD]) | 69 (4) | 68 (3.4) |
| Sex, Female (number [%]) | 7 (35.0) | 11 (57.9) |
| White (number [%]) | 20 (100.0) | 18 (94.7) |
| **Clinical** |  |  |
| BMI (kg/m<sup>2</sup>, mean [±SD]) | 26.1 (2.8) | 26.5 (4.3) |
| SBP (mm Hg, mean [±SD]) | 124.3 (13) | 121.1 (12.9) |
| **Laboratory** |  |  |
| Sodium (mEq/dL, mean [±SD]) | 137.7 (1.5) | 137.5 (1.6) |
| Hemoglobin (gm/dL, mean [±SD]) | 13.5 (0.9) | 13.1 (0.9) |
| eGFR MDRD, (mL/[min · 1.73m<sup>2</sup>], mean [±SD]) | 81.5 (14.7) | 82 (10.6) |
| **Medications** |  |  |
| Statins (number [%]) | 5 (25.0) | 2 (10.5) |

1 BMI=body mass index; SBP=systolic blood pressure; eGFR MDRD=estimated 2 glomerular filtration rate.

**Figure 1:**
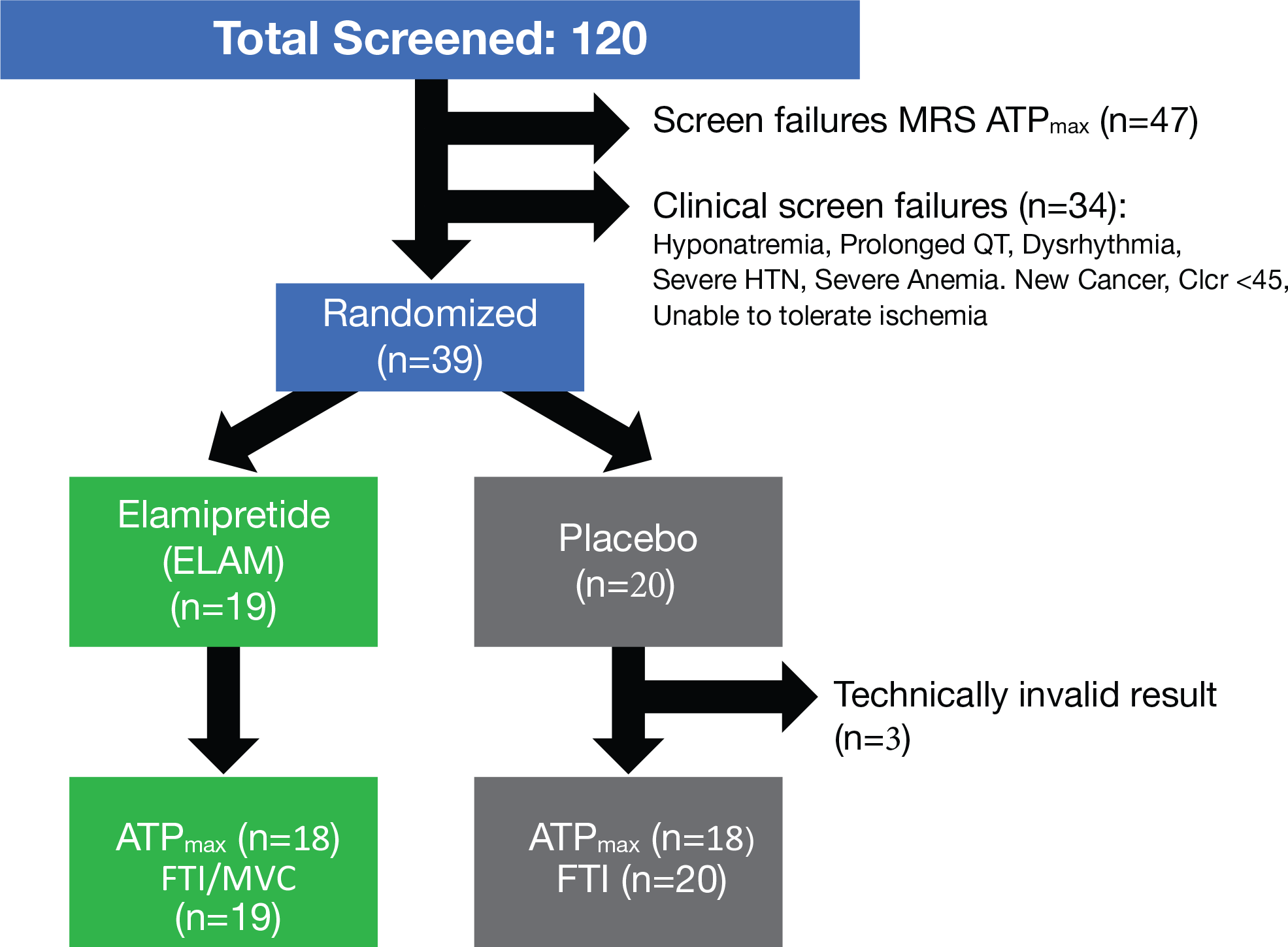
Study schema. The primary endpoint for the study was change in ATP_max_ by phosphorus magnetic resonance spectroscopy (^31^P MRS) after a 2-hour infusion compared with baseline (screening). The secondary endpoints were a change in mitochondrial coupling (P/O) and muscle force time integral (FTI) compared with baseline (screening).

### Elevated ATP_max_

The primary outcome of this study was an elevated ATP_max_ in the ELAM group versus the placebo group immediately after a 2-hour ELAM infusion (*P*=0.055 for delta ATPmax and P=0.045 for %change, **Figure 2A and B**). The values for ATPmax at baseline, immediate post-infusion, and 7 days after infusion, as well as the change values used in the ANCOVA analysis are reported in **Supplementary Table S3**. The mean increase from baseline to post-infusion was 27% in the in ELAM-treated subjects versus 12% in the placebo-treated subjects (**Figure 2B**). In the ELAM group both post-infusion and 7-day ATPmax values were elevated above baseline, while only the post-infusion ATPmax was higher than baseline in the placebo group (*P*<0.05, paired *t*-test). However, the change in ATPmax at 7 days was not different between ELAM and PL groups.

**Figure 2:**
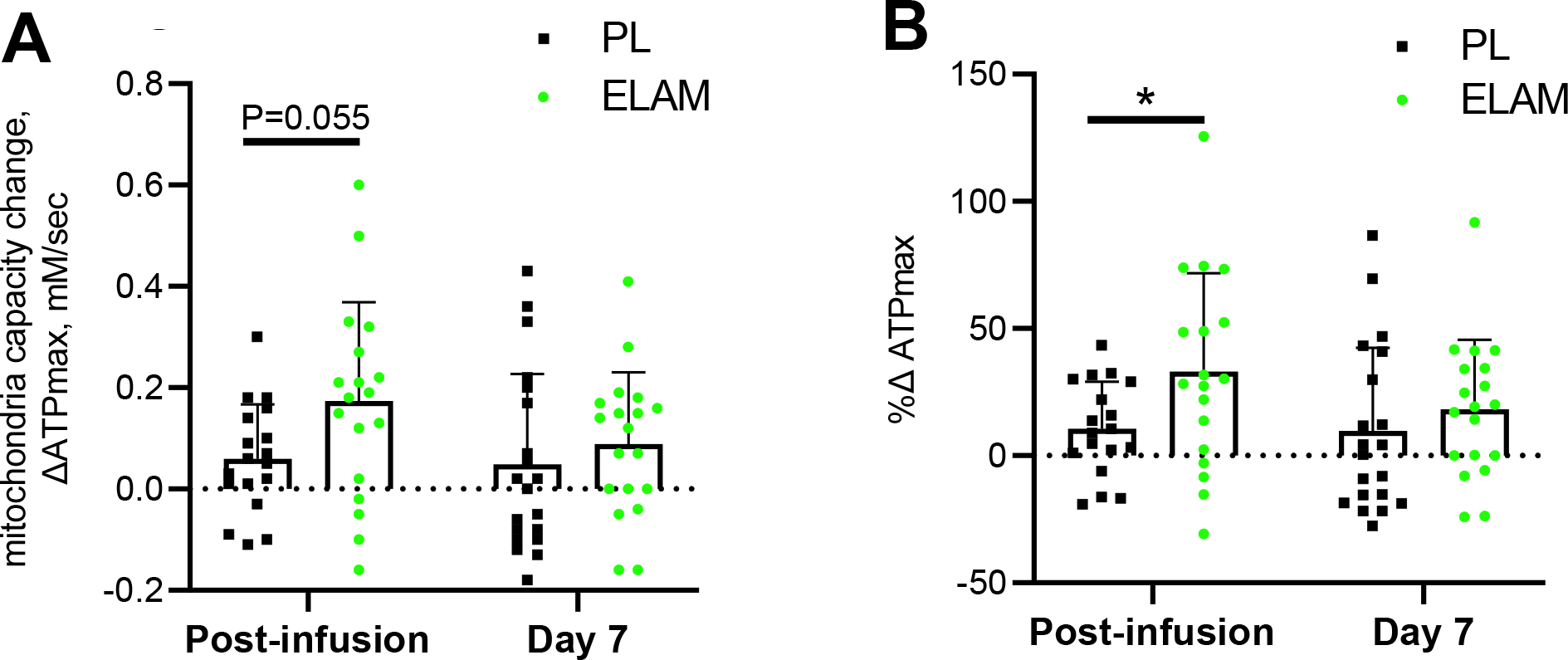
Change in mitochondrial energetic capacity (ΔATP_max_) with ELAM treatment. Change from baseline with ELAM treatment (post-infusion) and from baseline 7 days post-infusion (day 7). A) Absolute change in ATPmax; B) % change relative to baseline value. Error bars show SD. ELAM=elamipretide; PL=placebo.

### Resting P/O

A secondary endpoint was the efficiency of oxidative phosphorylation (mitochondrial coupling or P/O), which is one mechanism that could underlie the increased phosphorylation rate with ELAM treatment. No change was seen in this secondary outcome of resting P/O (**Figure 3**). The baseline, post-infusion and 7-day measurements and ANCOVA analyses are shown in **Supplementary Table S4**. This stability of resting P/O indicates that the improvement in ATP_max_ was the result of higher flux through the ETC rather than more efficient synthesis of ATP per O_2_ uptake. Thus, maximum flux through the ETC improved with ELAM treatment.

**Figure 3:**
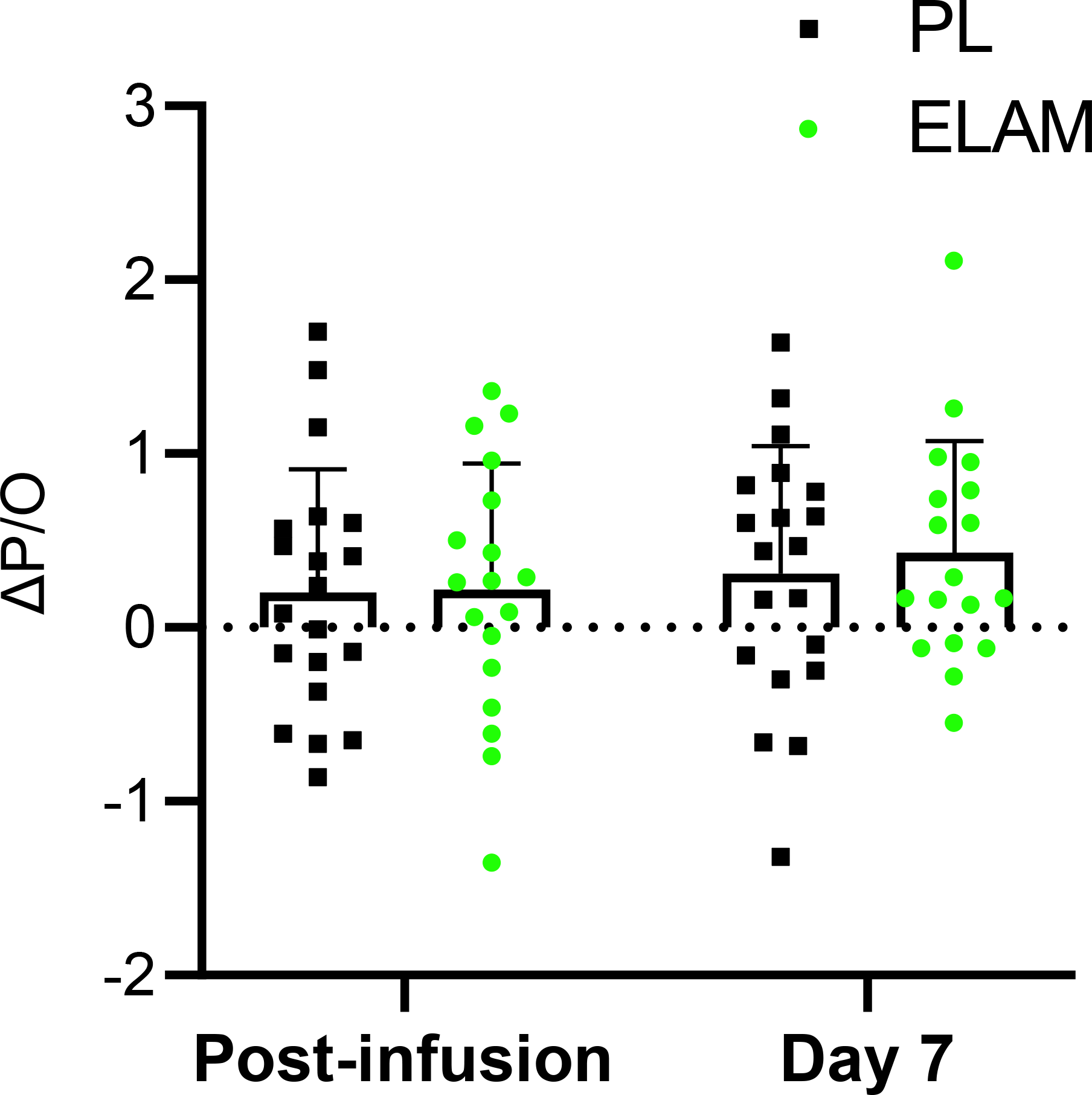
Change in mitochondrial coupling (ΔP/O) with ELAM treatment. Change from baseline with ELAM treatment (post-infusion) and from baseline 7 days post-infusion (day 7). Error bars show SD. ELAM=elamipretide; PL=placebo.

### Muscle Function

Exercise tolerance in the hand muscle was a secondary endpoint that assessed the impact of ELAM infusion on muscle function. This endpoint was measured as the sum of force produced during continuous isometric contractions to exhaustion (FTI) normalized to peak strength (FTI per maximum voluntary contraction [MVC]) [38]. No change was apparent in the FTI relative to placebo post-infusion with ELAM in the ANCOVA analysis specified in the protocol (Fig. 4; **Supplementary Table S5**). However, post-hoc analyses suggest that the study was not sufficiently powered to detect a difference in muscle function in the full ANCOVA model. The ELAM group significantly increased relative to baseline (*P*<0.033, paired t-test), while the placebo group did not (*P*=0.42). In addition, a significant rise was found with the two additional days of testing (day 1 and day 7 after infusion, *P*<0.04) between ELAM versus placebo in ANOVA. In addition, a post-hoc analysis defining muscle fatigue by the number of contractions performed instead of FTI reduced within group variability and indicated a significant treatment effect with a two-way ANOVA (**Figure S4**).

**Figure 4:**
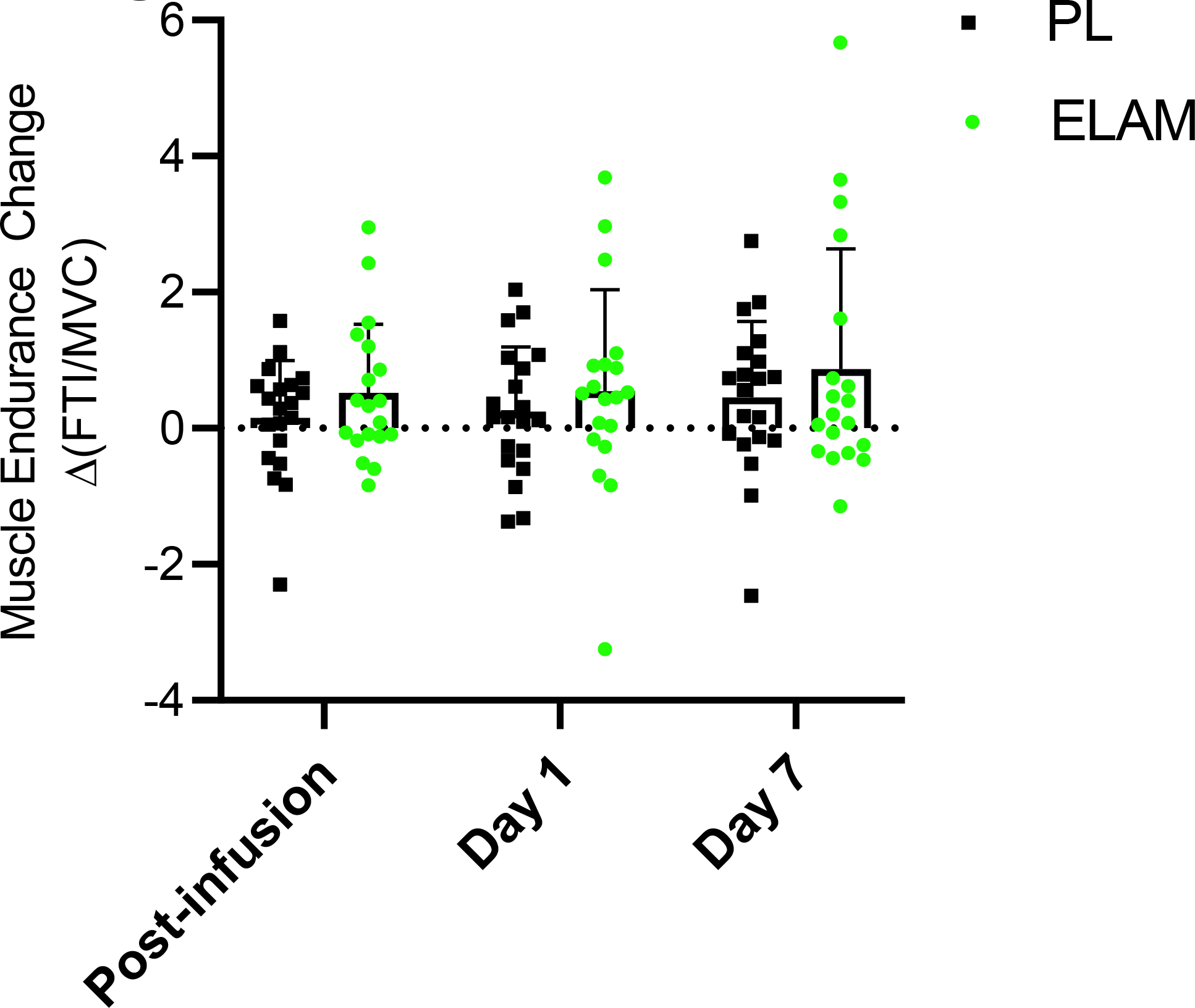
Change in muscle endurance test (ΔFTI/MVC) with ELAM treatment. Change from baseline with ELAM treatment post-infusion, from baseline 1 day post-infusion (day 1) and from baseline 7 days post-infusion (day 7). Error bars show SD. ELAM=elamipretide; PL=placebo.

## DISCUSSION

In a double-blind placebo-controlled randomized clinical trial a single 2-hour infusion of ELAM improves *in vivo* ATP_max_ in the FDI over placebo in skeletal muscle of older adults. Based on published materials that elucidated the mechanism of action of elamipretide, it has been postulated that this significant change in ATP_max_ can be attributed to the ability of elamipretide to reversibly bind to cardiolipin, resulting in stabilization of the inner mitochondrial membrane and optimization of the electron transport chain in dysfunctional mitochondria [39-41]. This rapid response of *in vivo* ATP_max_ to a single ELAM dose in older adult humans reproduces results observed in old mouse muscle using a similar MRS approach to measure mitochondrial function [14]. A new insight that arose from this study is that the rise in ATP_max_ on the day of treatment fades by day 7, which is consistent with the 16 hour half-time of ELAM in human blood [36]. Thus, ELAM acted to elevate mitochondrial ATP generation capacity in a single dose with a similar magnitude but much faster speed than prolonged interventions, such as exercise, in human muscle from older adults.

In this study we specifically selected community-dwelling older adult participants who demonstrated *in vivo* deficits in mitochondrial function to focus on the primary endpoint of the study a priori defined as the change in ATP_max_ with ELAM treatment. Our inclusion criteria included an ATP_max_ below 0.7 mM ATP/sec, which is above the mean ATP_max_ of 0.5 - 0.6 mM ATP/sec from quadriceps muscle we have observed in previous studies of older adults [9, 37, 42]. Previous work *in vivo* and *in vitro* suggests that some of this decline in oxidative capacity in the skeletal muscle of older adults is due to poor quality mitochondria as well as the loss of mitochondrial content with age. This is an important point because previous work in both mouse models and human subjects with genetic mitochondrial disease indicate that ELAM (SS-31) provides little or no benefit to well-functioning mitochondria [14, 26]. Despite the data described above, the extent and nature of mitochondrial dysfunction in aging skeletal muscle remains controversial with reports of no differences in oxidative capacity (ATP_max_) or maximum respiration in vitro between older and young adult subjects. The disparity in findings between studies underscores heterogeneity of mitochondrial function with aging helping to explain the observation that nearly 40% of older adults screened for the current trial were excluded due to normal functioning mitochondria.

Measurement of *in vivo* ATP_max_ provides a direct estimate of the capacity for mitochondrial phosphorylation, while other approaches use mitochondrial respiration as a surrogate for the capacity for ATP production [43]. Therefore, the increased ATP_max_ observed in this study and in previous work in aged mouse skeletal muscles could be due to: 1) increased capacity for flux through the electron transport system (ETS) and 2) improved coupling of oxidation (electron flux, oxygen consumption) to phosphorylation (ATP production) by the mitochondria (P/O) [14]. These two possibilities are not mutually exclusive and are both consistent with the hypothesis that ELAM reversibly binds to cardiolipin to stabilize the cristae structure in the inner mitochondrial membrane [23, 44]. ELAM (or SS-31) has been demonstrated to increase or protect maximum ETS flux in isolated mitochondria and permeabilized muscle fibers in the case of severe dysfunction in multiple models and human subjects associated with heart failure, ischemia-reperfusion, and disuse muscle atrophy. However, in aged mouse skeletal muscle the increased ATP_max_ after 1 hour or 8 weeks of ELAM (SS-31) treatment was not associated with increased state 3 respiration in permeabilized muscle fibers [14, 26]. It is worth noting that there was also no decrease in state 3 respiration comparing young to old in these studies despite the significant decline in ATP_max_ with age, suggesting there is a disconnect between maximum rates of respiration measured under saturating substrates and optimal conditions *ex vivo* and maximum ATP production measured under physiological conditions *in vivo*. In humans there is a significant correlation between ATP_max_ and state 3 respiration in permeabilized fibers and in young and old skeletal muscles, although *in vivo* ATP_max_ explained only about half of the variation in state 3 respiration in the older adult subjects. These results suggest increased maximum ETS flux may not fully explain the improved ATP_max_ with ELAM treatment. Recent work also indicates the ELAM directly interacts with mitochondrial proteins involved in substrate supply to the ETS and phosphorylation of ADP to ATP[45] raising the possibility that ELAM could both enhance substrate supply to the ETS and directly affect the phosphorylation system in aged mitochondria.

The lack of an effect of ELAM on the coupling of oxidative phosphorylation (P/O) at rest (**Supplemental Methods, Table S4**) in this study, despite selection of subjects with low P/O, indicates that changes in the P/O measured under resting conditions is not an important factor in the improved ATP_max_ observed in this study. The coupling between oxygen consumption and ATP production (P/O) decreases with age in mouse and human skeletal muscle [29]. In aged mice P/O increased with ELAM (SS-31) treatment 1 hour after a single dose and after 8 weeks of continuous treatment [14, 26]. It is important to note that *in vivo* P/O is measured under resting conditions where membrane potential is high, while ATP_max_ is measured under high flux conditions and lower membrane potential. Since membrane potential is a key driver of unregulated proton leak across the mitochondrial inner membrane and proton leak is an important mechanism underlying reduced coupling, the P/O is expected to increase as the flux through the ETS increases as has been demonstrated in isolated mitochondria [46]. Therefore, a lower P/O measured under resting conditions may not necessarily have a large impact on ATP production at the high ETS flux rates near ATP_max_. This argument and the lack of an effect of ELAM on P/O in this study suggests that mitochondrial coupling is not the primary driver of the increased ATP_max_ observed in this study. However, this does leave open the possibility that there is a defect in the phosphorylation system (e.g. ADP/ATP exchange, ATPase function) that impairs the coupling of respiration to ATP production as the mitochondria approach maximum capacity. If ELAM treatment led to reversal of hypothetical dysfunction in the ANT or ATPase this could have the effect of increasing ATP_max_ without a large effect on state 3 respiration. In support of this possibility is a new study using chemical cross linking with mass spectrometry (XL-MS) to demonstrate that ELAM directly interacts with both the ANT and F1F0 ATPsynthase in skeletal muscle mitochondria from aged mice [45]. However, the functional effects of these direct protein interactions remain to be tested.

The rapid increase in ATP_max_ after a 2-hour ELAM infusion agrees with the immediate effect of ELAM treatment on the same measure in the hindlimb of aged mice 1 hour after treatment [14]. Such a fast mitochondrial response is apparent in isolated organ models of disease after ELAM treatment [47]. In comparison, weeks to months are required to achieve a response with treatments that activate mitochondrial biosynthesis pathways. For example, an oral dose of a SIRT1 activator designed to activate mitochondrial biogenesis in older adult subjects involved a 21-day treatment and resulted in a trend toward improved oxidative phosphorylation capacity [48]. In contrast to building new mitochondria, the rapid response in ATP_max_ in old, but not in young muscle of mice, suggests that ELAM treatment repaired a dysfunction that quickly raised mitochondrial ATP supply [14]. A mechanism of action of ELAM that includes both direct protein interactions and association with cardiolipin to alter membrane structure resulting in improved ETC function and mitochondrial oxidative phosphorylation capacity is consistent with these findings.

Despite the effect on ATPmax, the rise in exercise tolerance did not achieve statistical significance in ELAM versus placebo groups on the infusion day (**Figure 4**, *P*=0.16 **Supplemental Table S5**). However, a significant rise was found between ELAM versus placebo with two additional days of testing (one day and one week post-infusion) in an ANOVA test (*P*<0.04). Both animal and human studies have demonstrated improvement in exercise tolerance with ELAM many days after infusion with daily doses. Greater 6MWT distance was found in patients with genetic mitochondrial disease and impaired mobility after 5 daily infusions at the dose used in this study [28]. Similarly, old mice showed increased endurance (running time to fatigue) on day 8 after daily treatment and after 8 weeks of continuous treatment [14].

This study was originally powered to detect a difference in the effect of treatment on mitochondrial energetics as the primary endpoint. As a result, the study was underpowered to detect an effect on muscle fatigue resistance based on the FTI/MVC measure. In a post-hoc analysis where the number of contractions, instead of FTI, was used to assess muscle fatigue the ELAM treatment increased muscle fatigue resistance on day 7 (*see supplemental material for methods and Figure S4*). The absence of significant improvement in muscle endurance despite the increased ATPmax immediately after infusion suggests a disconnect between mitochondrial ATP_max_ and muscle performance changes with ELAM treatment in older adult human muscle. The direct effects on the mitochondria occurring rapidly, while improvements in muscle performance may take longer to manifest and be due to downstream effects of mitochondrial function not directly linked to ATP supply by the mitochondria.

The primary limitation to this clinical trial was the small sample size target of 20 subjects per treatment group that was reduced to 18 per group with a complete ATPmax dataset due technical problems leading to missing data. This subject number achieved statistical significance for a relative ATP_max_ change (Δ%ATP_max_, *P*=0.045) with ELAM treatment, but 25 subjects are needed to reach the threshold for the absolute change in ATP_max_ based on power calculations derived from this study. A second limitation is the absence of data on *ex vivo* functional data and redox status from isolated mitochondria or permeabilized fibers. In the absence of the ability to specifically manipulate and interrogate specific components of oxidative phosphorylation that are only possible *ex vivo* we are left to speculate on the specific nature of the dysfunction with age and reversal with ELAM treatment observed *in vivo* in this study.

## CONCLUSIONS

Here, we demonstrate a pharmacological treatment that rapidly reverses mitochondrial deficits of ATP production in skeletal muscle in healthy older adult subjects. Several new insights come from this demonstration. First, the rise in ATP_max_ with a single infusion identifies the mitochondria as a site of action of ELAM. Second, the speed of the effect suggests that a rapidly reversible process in the mitochondria underlies this improvement. This is in contrast to the biosynthesis and replacement of mitochondrial components that underlie improvements in mitochondrial metabolism observed with exercise training. Third, the decline in ATP_max_ to placebo levels after one week indicates a reversible process. Thus, ELAM treatment provides new insight into the nature of mitochondrial and muscle dysfunction with age. Most importantly, ELAM treatment holds promise as a new therapy for the directly targeting mitochondrial dysfunction associated with age with the potential to improve mobility limitations and disability that come with age and disease.

## Data Availability

Data will be made available upon request.

## Acknowledgements

We thank the volunteers who participated in this study for their time and dedication as well as the contributions by Amy Good, Laura Ferrara, Jeff Finman, Peter Rabinovich, Doug Weaver, Marc Bamberger, and James Carr. The authors of this manuscript certify that they comply with the ethical guidelines for publishing in the Journal of Cachexia, Sarcopenia, and Muscle [49].

## Author contributions

Baback Roshanravan, David J. Marcinek, Kevin E. Conley, and Stealth BioTherapeutics designed the research; Kevin E. Conley, Sophia Z. Liu, Amir S. Ali, and Eric G. Shankland performed the research; Baback Roshanravan, John K. Amory, and Thomas Robertson provided clinical oversight; Chessa Goss coordinated the study; Kevin E. Conley, Sophia Z. Liu, David Marcinek, and Amir S. Ali analyzed data; and Kevin E. Conley, Baback Roshanravan, and David J. Marcinek wrote the paper. As first, second, and corresponding authors, Baback Roshanravan, Sophia Z. Liu, and David J. Marcinek contributed equally to the study. Kevin E. Conley contributed to the manuscript, but died prior to the manuscript’s submission for publication.

