## Supplemental information for "*In Vivo* Mitochondrial ATP Production Is Improved in Older Adult Skeletal Muscle After a Single Dose of Elamipretide in a Randomized Trial"

| **Table S1: Inclusion Criteria** |
| --- |
| 1. Are male and female adults aged ≥60 and ≤85 years |
| 2. Have *in vivo* $^{31}$P MRS and OS determined $ATP_{max}$ < 0.70 mM/sec |
| 3. Have *in vivo* $^{31}$P MRS and OS determined P/O < 1.9 |
| 4. Are ambulatory and able to perform activities of daily living without assistance |
| 5. Have sufficient venous access for study drug administration and clinical testing |
| 6. Speak and read English fluently |
| 7. Provide informed consent |

Abbreviations - $^{31}$P MRS: magnetic resonance spectroscopy; OS: optical spectroscopy; ATPmax: maximum rate of oxidative phosphorylation.

| Table S2. Exclusion Criteria |
|---|
| 1. Have significant disease(s) or condition(s) that put the subject at risk |
| 2. Rhabdomyolysis |
| 3. Hospitalized within 3 months for major atherosclerotic events |
| 4. Any metal implants in soft tissues |
| 5. Implanted cardiac pacemaker or other implanted cardiac device |
| 6. Serum sodium level < 136 mEq/L at Screening or Pre-infusion |
| 7. Hemoglobin level < 12 g/dL at Screening or Pre-infusion |
| 8. Have chronic, uncontrolled hypertension as judged by the Investigator |
| 9. BMI of < 18 or > 32 $kg/m^2$ |
| 10. Creatinine clearance < 45 mL/min |
| 11. Laboratory or ECG abnormalities |
| 12. Endocrine disorder or Peripheral neuropathy |
| 13. Have a history of autoimmune disease (e.g., lupus, rheumatoid arthritis) |
| 14. Difficulty using subject's right hand |
| 15. Claustrophobia |
| 16. Cancer, unless subject has documentation of completed curative treatment |
| 17. History of or risk factors for deep vein thrombosis or pulmonary embolism |
| 18. History of serious mental illness as judged by the Investigator |
| 19. Body temperature > 37.5°C at the time of planned dosing |
| 20. Alcohol or drug abuse |
| 21. Donated or received blood or blood products within the past 30 days |
| 22. Clinically significant abnormal 12-lead ECG |
| 24. Directly affiliated with this study, sponsor employees and/or their immediate families. |
| 25. Are currently enrolled or have participated, within the last 30 days in a clinical study involving an investigational product including Elamipretide |

**Table S3.** $ATP_{max}$ ($mM\ sec^{-1}$, primary endpoint) post-infusion and 7 days later compared to baseline.

| **post-infusion** | **Baseline** | **immediate** | **Change** | ***P*** |
|---|---|---|---|---|
| Placebo | 0.56 (0.08) | 0.63 (0.14) | 0.06 (0.11) | 0.055 |
| ELAM | 0.59 (0.13) | 0.75 (0.21) | 0.17 (0.20) | |
| **7 days post-infusion** | **Baseline** | **7 day** | **Change** | ***P*** |
| Placebo | 0.56 (0.07) | 0.61 (0.18) | 0.05 (0.18) | 0.283 |
| ELAM | 0.59 (0.13) | 0.68 (0.15) | 0.09 (0.14) | |

***P*** - ELAM vs. PL in an ANCOVA. Mean (SD).

**Table S4.** P/O (mitochondrial coupling, secondary endpoint) post-infusion and 7 days later compared to baseline.

| **post-infusion** | **Baseline** | **immediate** | **Change** | ***P*** |
|---|---|---|---|---|
| Placebo | 1.47 (0.39) | 1.73 (0.68) | 0.26 (0.69) | 0.98 |
| ELAM | 1.46 (0.33) | 1.72 (0.60) | 0.25 (0.68) | |
| **7 days post-infusion** | **Baseline** | **7 day** | **Change** | ***P*** |
| Placebo | 1.47 (0.39) | 1.97 (0.73) | 0.51 (0.79) | 0.36 |
| ELAM | 1.46 (0.33) | 1.79 (0.52) | 0.32 (0.47) | |

***P*** - ***P*** - ELAM vs. PL in an ANCOVA. Mean (SD).

**Table S5.** FTI (secondary endpoint) post-infusion, 1 day and 7 days later compared to baseline.

| **post-infusion** | **Baseline** | **immediate** | **Change** | ***P*** |
|---|---|---|---|---|
| Placebo | 3.43 (1.31) | 3.53 (1.34) | 0.10 (0.73) | 0.16 |
| ELAM | 3.48 (1.23) | 3.91 (1.13) | 0.43 (0.88) | |
| **1 day post-infusion** | **Baseline** | **1 day** | **Change** | ***P*** |
| Placebo | 3.43 (1.31) | 3.67 (1.78) | 0.24 (0.95) | 0.49 |
| ELAM | 3.48 (1.23) | 3.95 (1.21) | 0.46 (1.04) | |
| **7 days post-infusion** | **Baseline** | **7 day** | **Change** | ***P*** |
| Placebo | 3.43 (1.31) | 3.87 (1.81) | 0.44 (1.11) | 0.35 |
| ELAM | 3.48 (1.23) | 4.31 (1.57) | 0.83 (1.48) | |

***P*** - ELAM vs. PL in an ANCOVA. Mean (SD).

**Figure S1. Determination of mitochondrial phosphorylation capacity from [PCr] recovery following brief exercise.** The continuous line (red) is the monoexponential fit to the data for a subject in the study (same as in Fig. S2). The vertical dashed line denotes the time constant for the PCr recovery fit to a monoexponential. The mitochondrial phosphorylation capacity is determined as: ATPmax = [PCr]/$\tau$, where [PCr] = 24.5 mM and $\tau$= 38 sec.

**Figure S2. Spectroscopic measurements during ischemia that yield $O_2$ uptake, ATP flux, and energy coupling (P/O) in an FDI muscle from a study participant at rest.** (Upper panel) Spectroscopic measurement of the change in Mb-$O_2$ and Hb-$O_2$ % saturation from onset of ischemia to 50% Mb-$O_2$ saturation, which is the range over which mitochondrial $O_2$ uptake is not $O_2$ limited. $O_2$ uptake is determined from the in vivo Mb and Hb concentrations times the % change in $O_2$ saturation. (Lower panel) MRS measurement of the [PCr] change in the ischemic muscle. This [PCr] breakdown measures the ATP turnover from the cell that is required from the mitochondria under aerobic conditions.

**Figure S3**: **Measurement of muscle force.** *A)* Apparatus for measuring force and contraction number in the hand muscle, first dorsal interosseous. Pictured is the magnetic resonance coil located over the hand muscle to collect the $^{31}P$ magnetic resonance spectral measurements that determined $ATP_{max}$. *B)* Force measurements used to determine the maximum voluntary contraction (MVC) and force-time integral (FTI) to fatigue. The single force records generated by the hand muscle were used to determine MVC. The force measurement during repeated contractions to fatigue are shown. These contraction records were summed into the FTI. The red tracings show an example of the sum of the force generation during contraction at baseline (pre-treatment). The blue tracings show a greater number of contractions and summed FTI in the exercise test 7 days post-infusion. The vertical arrows designate the point of fatigue.

**Figure S4**: Post-hoc analysis of muscle performance changes with ELAM treatment. A) Change in the number of contractions completed at fatigue from baseline with ELAM infusion (V2-V1) and on day 7 after infusion (V4-V1; mean±SEM). 2-way ANOVA indicates a significant treatment effect (P=0.027). P values in figure are multiple comparisons test.

**Figure S1**

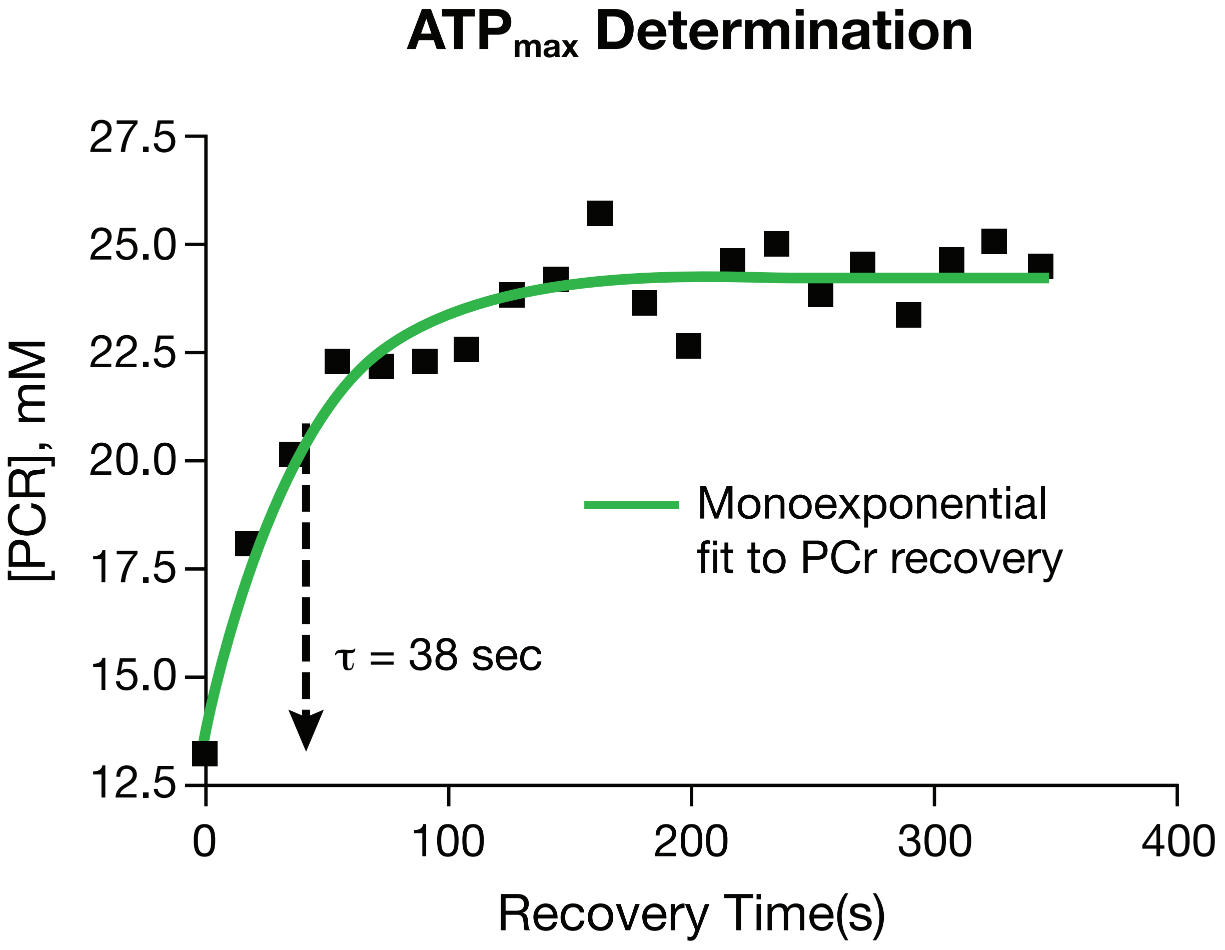

ATP$_{max}$ Determination
27.5
25.0
22.5
20.0
17.5
15.0
12.5
[PCR], mM
0
100
200
300
400
Recovery Time(s)
Monoexponential fit to PCr recovery
τ = 38 sec

### Figure S2

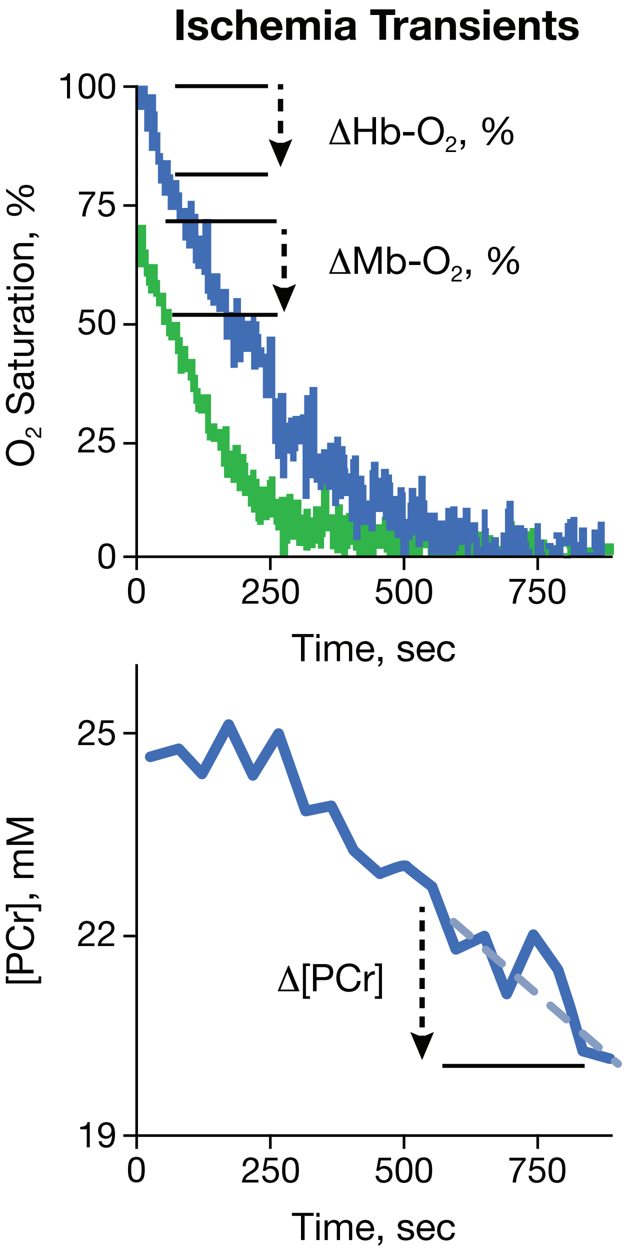
Ischemia Transients
ΔHb-O2, %
ΔMb-O2, %
O2 Saturation, %
100
75
50
25
0
0
250
500
750
Time, sec
[PCr], mM
25
22
19
Δ[PCr]
0
250
500
750
Time, sec

### Figure S3

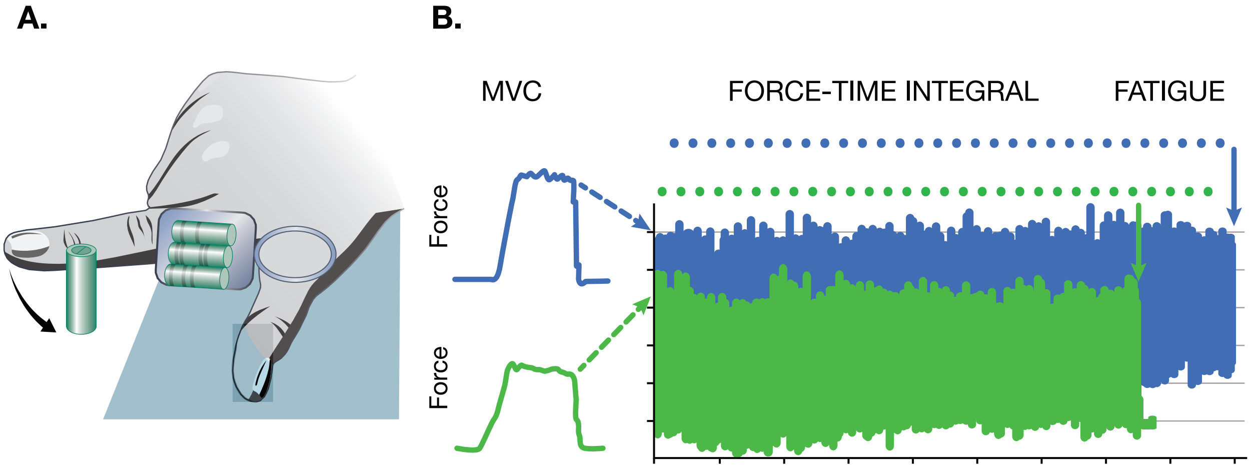

#### Figure S4

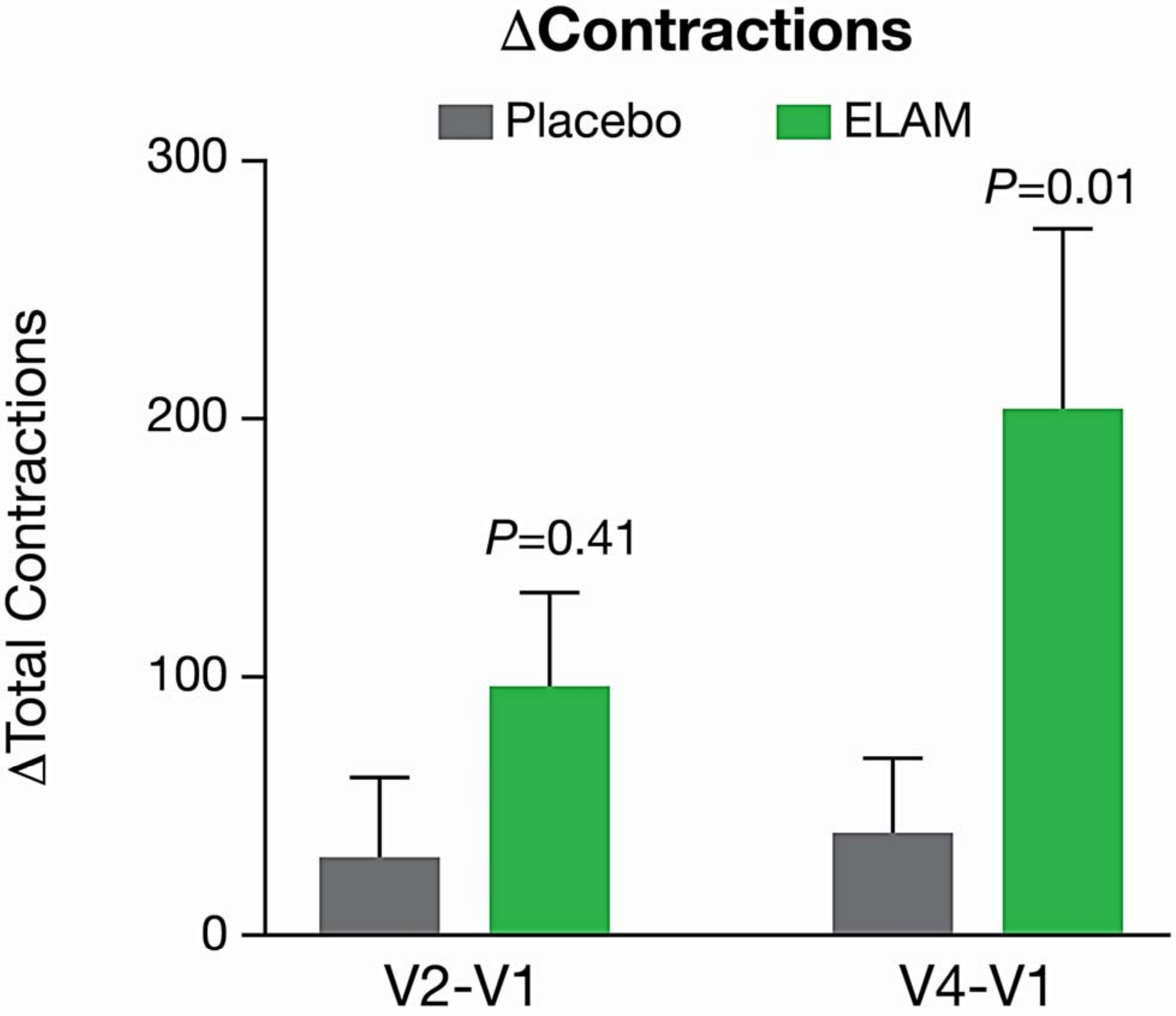
ΔContractions
Placebo
ELAM
300
200
100
0
ΔTotal Contractions
P=0.41
P=0.01
V2-V1
V4-V1
